# A psychometric validation of the Hospital Anxiety and Depression Scale (HADS) in community-dwelling older adults

**DOI:** 10.1101/2022.07.08.22277407

**Authors:** Heidi Emly Sivertsen, Anne-Sofie Helvik, Linda Gjøra, Gørill Haugan

## Abstract

**Objectives:** The Hospital Anxiety and Depression scale (HADS) is commonly used to measure anxiety and depression, but the number of studies validating psychometric properties in older adults are limited. To our knowledge, no previous studies have utilized confirmative factor analyses in community-dwelling older adults, regardless of health conditions. Thus, this study aimed to examine the psychometric properties of HADS in older adults living at home in a large Norwegian city.

**Methods:** In total, 1190 inhabitants ≥70 (range 70 – 96) years completed the HADS inventory in the population-based Trøndelag Health Study (HUNT); termed “HUNT4 70+” in Trondheim, Norway. Confirmatory factor analyses (CFA) were performed to test the dimensionality, reliability, and construct validity.

**Results:** The original two-factor-solution (Model-1) revealed only partly a good fit to the present data; however, including a cross-loading for item 6_D_ (*“I feel cheerful”*) along with a correlated error term between item 2_D_ (*“I still enjoy the things I used to enjoy”*) and 12_D_ (*“I look forward with enjoyment to things*”), Model-3 with a two-factor solution demonstrated an acceptable/good fit. Good to acceptable measurement reliability was demonstrated, and construct validity was supported.

**Conclusions:** The HADS involves some items which are not reliable and valid indicators for the depression construct in this population. Especially item 6 which is problematic. To improve the reliability and validity of HADS rewording of some items is recommended.

## Introduction

Older adults (65+) living at home may have multiple diseases, impaired physical function (1-6) and impaired cognitive function (6-8). Compared with younger age groups, less characteristic symptoms of depression often appear in old ages, and depression may be detected and diagnosed less frequently among older adults (3, 8-12). In addition, many community-dwelling older adults who meet diagnostic criteria for depression do not seek health care for their symptoms; less than half have contact with the health service and barely 10 % receives effective treatment (8). However, major depressive disorder (MDD) and clinically significant depressive symptoms (CSDS) are associated with decreased quality of life (QOL), increased comorbidity with physical illness, reduced physical functioning in daily activities, increased risk of dementia (13) and increased need for help and risk of mortality (7, 10, 11, 14).

Depression is prevalent among older adults worldwide with an estimate of depressive disorders of 5.4%, including MDD and CSDS (aged >70 years) (15). Furthermore, a recent review based on 20 studies found the pooled prevalence of MDD to be 13.3% (16). The pooled prevalence of CSDS has been reported to be higher than for MDD. Two recent systematic reviews, largely based on studies of community-dwelling older adults, found the pooled prevalence to be 28.4% and 31.7%, respectively. Moreover, these two reviews found a large variety of the prevalence rates ranging between 7.7 – 81.1%; the authors explained this variety by cultural differences, along with differences in sample characteristics, methodology, and screening tools (17, 18).

Depression often co-exists with anxiety and anxiety symptoms. Compared to younger age groups, older adults report a higher frequency of anxiety (8, 19). In 2019, the estimated prevalence of anxiety disorders (a combined estimate of all subtypes) among adults aged >70 years was 4.4% (15). A recent review among community-dwelling adults > 55 years old revealed a pooled prevalence of anxiety disorders and anxiety symptoms of 5.4% and 7.9%, respectively (20). Generalized anxiety disorder is the most prevalent anxiety disorder and leads to high mortality risk, which is even higher when accompanied by depression (21).

Similar to MDD and CSDS and compared to younger age groups, older adults report less characteristic symptoms of anxiety disorders and anxiety symptoms (20); for instance are worries about health, disturbances of sleep and less reassurance seeking behaviors common symptoms of anxiety among older adults (19). Consequently, anxiety disorder and anxiety symptoms may be detected and diagnosed less frequently among older adults. It is also known that older adults are less likely to seek health care for their symptoms than younger adults, because older adults are less likely to report their symptoms, have less knowledge regarding anxiety disorders and available treatments, with barriers to treatment seeking which include stigma, cost, transportation, and mobility (22).

Another explanation for this could be that common instruments are developed for younger age groups and might be less suitable for older adults (20).

The Hospital Anxiety and Depression Scale (HADS) is a screening tool commonly used in epidemiological research to estimate the prevalence of anxiety symptoms and CSDS among older adults (17). Originally, the HADS aimed to measure states of anxiety and depression in adults receiving treatment for physical health problems in general hospitals (23). HADS consists of 14 items and includes subscales for anxiety (HADS-A; seven items) and depression (HADS-D; seven items). The items are scored on a four-point scale ranging from totally disagree to totally agree. Each item is rated from 0-3, where higher scores indicate more anxiety and/or depression. The maximum score on each subscale is 21 ranging from 0-7 (normal), 8-10 (mild disorder), 11-14 (moderate disorder), to 15-21 (severe disorder) (23). HADS has been widely used (24, 25), suggested with a lower cut-off for older adults (26) and tested with satisfactory psychometric properties (24). However, to a less extent among older adults.

To increase acceptability and avoid individuals feeling as though they are being tested for mental disorders, symptoms of severe psychopathology have been excluded, this makes HADS more sensitive to milder psychopathology (27, 28). According to the International Classification of Diseases (ICD-10) five of seven items in HADS-D focus on lack of positive feelings and cover only two of three main criteria for depression; physical symptoms as loss of energy, sleep- and appetite disturbances are not covered.

The Norwegian version of the HADS is psychometrically evaluated among older adults without cognitive impairment in nursing homes (28), older adults admitted to somatic hospitals (29), older community-dwelling adults (30) and among adults consulting the general practice (31). However, with one exception (28) Norwegian studies have evaluated the psychometrical features of HADS by means of principal component analysis (PCA) (27, 29, 31) without utilizing the confirmative approach. Internationally, a few studies have studied features of HADS using confirmative factor analysis (CFA) in community-dwelling, non-clinical samples of older adults (24, 32, 33). These studies give support to the original two-factor structure showing good internal consistency. Even so, problems with cross-loadings and ceiling effect are reported (24). Utilizing CFA a psychometrical evaluation of HADS in a clinical sample of veterans reported a three-factor structure to fit better than the original two-factor solution (34).

Up to date, the factor structure, the internal consistency, construct validity and homogeneity of the HADS have not been assessed by means of CFA among community-dwelling older adults ≥70 years in Norway. Therefore, the aim of the present study was to examine the psychometric properties of the HADS scale among community-dwelling older adults in Norway by investigating its dimensionality, reliability, and construct validity. *Dimensionality* concerns the homogeneity of a scale’s items (35), indicating if the included items match the defined construct. Depression and anxiety have been seen to correlate strongly but are still considered different constructs. *Reliability* encompasses a scale’s consistency and lack of error (36). To assess internal consistence of the items the reliability coefficients of Cronbach’s alpha (α) and composite reliability (ρ_c_) were utilized. Finally, *construct validity* implies various aspects such as convergent, discriminant and content validity. In this study, convergent and discriminant validity denotes if HADS relates with other constructs as expected, while content validity embraces whether the theoretical content of the Anxiety and Depression constructs involved in HADS are adequately represented by the 14 items. This is, whether the included items cover the theoretical definition they are aimed to represent (37). If the wording of items is too similar, Cronbach’s alpha, content validity, and dimensionality will be falsely improved. In consequence, the average correlation among items increases, and therefore also coefficient alpha; however, without adding substantially to the content validity of the scale. Obviously, to tap into the same construct some similarity/correlation is needed. Nevertheless, items simply representing a rephrasing of other items are redundant.

## Methods

### Sample and procedures

During 2017-2019 persons aged 70+ were recruited from the city of Trondheim, county of Trøndelag in Central Norway as a part of the fourth wave of The Trøndelag Health Study (HUNT). HUNT is a population-based cross-sectional study (38). The “HUNT 4 70+ Study” includes the entire population in the county; 9930 (64%) consented to participate (39). The HUNT study comprises questionnaires, clinical measurements, and collections of biological samples.

Totally, 5087 inhabitants in the city of Trondheim were invited to participate. The inclusion criteria were age of ≥ 70 years and permanent residency in two randomly selected districts of the city. Out of the 5078, a total of 1747 (response rate of 34.3%) individuals participated (57.9% women and 42.1% men). The participants underwent a comprehensive clinical evaluation at the field-station and received a visit at home (some lived in a nursing home). In addition, they filled in two questionnaires (Q1 and Q2), either at the field station or at home. The questionnaire was sent to the field station by mail. HADS was included in Q2.

In this study, the inclusion criterion was to be community-living, while the exclusion criteria included: (1) lack of responses in HADS (n=392); (2) nursing home residents (n=65); and (3) lack of other essential health variables (n=100 participants). In total, 557 were excluded leaving an effective sample comprising 1190 participants.

### Statistical analysis

The analysis were conducted using the IBM Statistical Package for the Social Sciences (SPSS) Version 28 software (40) and Stata 17 software package (41). Confirmatory Factor Analysis (CFA) represents a more accurate evaluation of the psychometric properties of scales used. In this study, the model fit adequacy was assessed by χ^2^-statistics and conventional fit indices: χ^2^-statistics, the Root Mean Square Error of Approximation (RMSEA) and the Standardized Root Mean Square Residual (SRMR) with values <0.10 are acceptable and values ≤0.05 indicates a good fit (42, 43). Further, the Comparative Fit Index (CFI) and the Tucker-Lewis Index (TLI) with acceptable fit at 0.95 and good fit at 0.97 (42-45). Skewness and kurtosis were significant. Therefore, the Satorra-Bentler corrected χ^2^ which is the correct asymptotic mean even under non-normality is reported (46).

## Results

### Sample characteristics

The sample of 1190 adults were aged between 70-96 years with a mean of 76.5 (SD = 5.3); 644 (54.1%) were women (mean age 76.7) while 546 (45.9%) were men (mean age 76.2) (Table 1). Furthermore, 325 (27.3%) had completed higher education (≥4 years of university/college). About 50% had physical or mental long-term illness, injury, or a disorder that impaired their daily functioning (47.1%), only a few had any kind of in-home care (4.7 %), nursing care (4.5 %) and/or had been admitted to a nursing home for a period during the last year (3.3%).

**Table 1.** Demographic characteristics of the study population by gender.

|  | Total<br>(N=1190) | Women<br>(n= 644, 54%) | Men<br>(n=546, 46%) | p-value |
| --- | --- | --- | --- | --- |
| Age, M (SD) | 76.5 (5.3) | 76.7 (5.4) | 76.2 (5.1) | 0.004<sup>a</sup> |
| Married, n (%) | 709 (59.5) | 299 (25.2) | 410 (34.5) | < 0.001<sup>b</sup> |
| Education n (%) |  |  |  | < 0.001<sup>b</sup> |
| Primary school | 115 (9.7) | 87 (13.5) | 28 (5.1) |  |
| High School 1-2 years | 190 (16) | 138 (21.4) | 52 (9.5) |  |
| High School 3 years | 125 (10.5) | 77 (12) | 48 (8.8) |  |
| Apprenticeship certificate | 132 (11.1) | 56 (8.7) | 76 (13.9) |  |
| University/college < 4 years | 303 (25.5) | 148 (23) | 155 (28.4) |  |
| University/college > 4 years | 325 (27.3) | 138 (21.4) | 187 (34.2) |  |
| Morbidity, n (%) |  |  |  |  |
| Asthma | 125 (10.5) | 84 (13) | 41 (7.5) | 0.001<sup>b</sup> |
| Diabetes | 92 (7.7) | 39 (6.1) | 53 (9.7) | < 0.005<sup>b</sup> |
| Dementia | 75 (6.3) | 39 (6.1) | 36 (6.5) | n.s. |
| Heart attack | 102 (8.6) | 36 (5.6) | 66 (12.1) | < 0.001<sup>b</sup> |
| Impaired function in daily life<br>> one year | 560 (47.1) | 325 (58) | 235 (42) | < 0.005 |
| Home care and hospitalized, n (%) |  |  |  |  |
| Home care last year | 56 (4.7) | 37 (5.7) | 19 (3.5) | n.s.<sup>b</sup> |
| Nursing care last year | 53 (4.5) | 35 (5.4) | 18 (3.3) | n.s.<sup>b</sup> |
| Hospitalized nursing home last year | 39 (3.3) | 24 (3.7) | 15 (2.7) | n.s<sup>b</sup> |
| Overall global QOL | 1172 (98.5) | 636 (98.7) | 536 (98.2) | < 0.001<sup>b</sup> |

Note. HADS (Hospital Anxiety and Depression Scale). <sup>a</sup> Independent sample t-test, <sup>b</sup> Pearson chi-square test. n.s = not significant.

Looking at those who were excluded, these were significantly older (mean age 83.3; SD=8.0 years), more often female (65.7%) and less high education (≥4 years of university/college; 7.2 %). Dementia diagnosis was also more prevalent among the excluded participants (56.2% versus 6.3%).

### HADS item score statistics

The mean anxiety and depression scores were 3.4 (SD = 2.9) and 3.0 (SD = 2.5), respectively (Table 2.) The internal consistence of the anxiety and depression constructs (Table 2) was good (α_anxiety_=.79=) or acceptable (α_depression_=.66). Composite reliability (***ρ***c) displayed values between 0.65-0.92 (Table 3); values ≥0.60 are acceptable, whereas values ≥0.70 are good (42, 47).

**Table 2.** Means, Standard deviation (SD) and Cronbach’s alpha for the Norwegian version of the Hospital Anxiety and Depression Scale (HADS).

| Items | Response <sup>a</sup> |  |  |  | Total | Mean | SD | Cronbach's alpha (α) |
| --- | --- | --- | --- | --- | --- | --- | --- | --- |
|  | 0 | 1 | 2 | 3 |  |  |  |  |
| 1<sup>A</sup> I feel tense or 'wound up'* | 911 | 245 | 28 | 6 | 1190 | .27 | .52 |  |
| 2<sup>D</sup> I still enjoy the things I used to enjoy | 704 | 450 | 28 | 8 | 1190 | .45 | .58 |  |
| 3<sup>A</sup> I get a sort of frightened feeling as if something awful is about to happen* | 641 | 370 | 130 | 49 | 1190 | .65 | .83 |  |
| 4<sup>D</sup> I can laugh and see the funny side of things | 924 | 236 | 27 | 3 | 1190 | .25 | .50 |  |
| 5<sup>A</sup> Worrying thoughts go through my mind* | 778 | 299 | 90 | 23 | 1190 | .46 | .72 |  |
| 6<sup>D</sup> I feel cheerful* | 828 | 302 | 56 | 4 | 1190 | .36 | .59 |  |
| 7<sup>A</sup> I can sit at ease and feel relaxed | 661 | 480 | 48 | 1 | 1190 | .49 | .58 |  |
| 8<sup>D</sup> I feel as if I'm slowed down* | 382 | 668 | 99 | 41 | 1190 | .83 | .72 |  |
| 9<sup>A</sup> I get a sort of frightened feeling like 'butterflies' in the stomach | 702 | 464 | 21 | 3 | 1190 | .43 | .54 |  |
| 10<sup>D</sup> I have lost interest in my appearance* | 860 | 255 | 50 | 25 | 1190 | .36 | .66 |  |
| 11<sup>A</sup> I feel restless as if I must be on the move* | 451 | 568 | 152 | 19 | 1190 | .78 | .72 |  |
| 12<sup>D</sup> I look forward with enjoyment to things | 733 | 338 | 107 | 12 | 1190 | .49 | .70 |  |
| 13<sup>A</sup> I get sudden feelings of panic* | 909 | 246 | 26 | 9 | 1190 | .27 | .54 |  |
| 14<sup>D</sup> I can enjoy a good book or radio/TV program | 990 | 167 | 22 | 11 | 1190 | .21 | .51 |  |
| Total A |  |  |  |  | 1190 | 3.35 | 2.9 | .79 |
| Total D |  |  |  |  | 1190 | 2.95 | 2.5 | .66 |
| Total A+D |  |  |  |  | 1190 | 6.30 | 4.6 | .80 |

Note. N=1190. A= anxiety and D= depression. <sup>a</sup> Items were scored on a four-point scale ranging from totally disagree to totally agree. A-response: 0= 'Not at all', 1= 'Not very often', 2= 'Quite often' or 3= 'Very often' and D-response: 0= 'Most of the time', 1= 'Sometimes', 2= 'Not often' or 3= 'Not at all'. N= 1190. The standard scoring algorithm was used for A = sum of items 1\*, 3\*, 5\*, 7, 9, 11\*, 13\*; and for D = sum of items 2, 4, 6\*, 8\*, 10\*, 12, 14. \* Items starred are reverse scored.

**Table 3.** Goodness-of-fit indices for HADS measurement models: Model-1^1^, Model-2^2^, Model-3^3^.

| Fit Measure | Model-1<br>2-factors | Model-2<br>2-factors | Model-3<br>2-factors | Model<br>Anxiety | Model Depression |
| --- | --- | --- | --- | --- | --- |
| $\chi^2$ Satorra Bentler | 395.010 | 317.951 | 296.919 | 61.695 | 84.264 |
| p-value | <0.0001 | <0.0001 | <0.0001 | <0.0001 | <0.0001 |
| $\frac{\chi^2}{df}$ Satorra Bentler | 5.20 | 4.18 | 4.01 | 4.41 | 4.52 |
|  | Df=76 | Df=75 | Df=74 | Df=14 | Df=14 |
| RMSEA | 0.059 | 0.052 | 0.050 | 0.054 | 0.065 |

|  | (CI: 0.054-0.065) | (CI: 0.046-0.058) | (CI: 0.044-0.056) | (CI: 0.040-0.068) | (CI: 0.052-0.079) |
| --- | --- | --- | --- | --- | --- |
| p-value (*close fit test*) | 0.004 | 0.262 | 0.452 | 0.312 | 0.030 |
| SRMR | 0.052 | 0.042 | 0.040 | 0.029 | 0.38 |
| CFI | 0.91 | 0.93 | 0.94 | 0.97 | 0.94 |
| TLI | 0.89 | 0.92 | 0.92 | 0.96 | 0.92 |
| <sup>4</sup>Composite Reliability | $\rho_{\text{anxiety}}$ 0.78 | 0.78 | 0.78 | 0.78 | - |
| | $\rho_{\text{depression}}$ 0.70 | 0.68 | 0.65 | - | 0.70 |

Note. N=1190. <sup>1</sup>**Model-1**=original 2-factor-model with 14 items. <sup>2</sup>**Model-2**=Model-1 including a path from item 6<sub>D</sub> to ANXIETY. <sup>3</sup>**Model-3**=Model-2 including a correlated error between item 2<sub>D</sub> and 12<sub>D</sub>. RMSEA=Root Mean Square Error of Approximation. SRMR=Standardized Root Mean Square Residual. CFI=Comparative Fit Index. TLI=Tucker-Lewis Index.

### Confirmatory Factor Analysis (CFA)

We tested the original two-factor solution including 14 items and termed this solution **Model-1**; the factor loadings (λ) ranged between 0.32 and 0.72, followed by multiple squared correlations (R^2^) from 0.10 to 0.52. Three items belonging to the depression construct; 8_D_ (“I feel as I’m slowed down”), 10_D_ (“I have lost interest in my appearance”) and 14_D_ (“I can enjoy a good book or a TV program”) revealed low loadings 0.34, 0.32 and 0.37, respectively; explaining 12%, 11% and 13% of the variance of the depression construct. The fit indices indicated misspecification: χ^2^=395.010, p=0.00001, df=76, χ^2^/df=5.20, RMSEA=0.059, p-close=0.04, SRMR=0.052, CFI=0.91, TLI=0.89 (Table 3). The RMSEA, which is an estimate of approximate fit was acceptable, while the χ^2^ was much too high. For an acceptable fit the χ^2^/df should be ≤3.0, and ≤2 for a good fit. Further, the CFI and TLI were too low, all of which indicating misspecification. Exploring the normalized residuals, 23 residuals were significant with item 6_D_ (*“I feel cheerful”*) involved in several highly significant estimates. Hence, we scrutinized the modification indices (MI) presenting some extremely high values; item 6_D_ exposed an extremely high MI=77.29 with the anxiety factor, and an MI=30.374 with item 5_A_ (“*Worrying thoughts go through my mind*”). Also, item 2_D_ (*“I still enjoy the things I used to enjoy”*) and 12_D_ (*“I look forward with enjoyment to things*”) demonstrated an exceptionally high MI=44.383. In total, 16 MIs were ≥10.

Looking at one factor at a time, both anxiety and depression including 7 items each demonstrated a too high χ^2^, while the other indices were good to acceptable (<u>*Anxiety*</u>: χ^2^=61.695, p=0.00001, df=14, χ^2^/df=4.41, RMSEA=0.054, p-close=0.320, SRMR=0.029, CFI=0.97, TLI=0.96; <u>*Depression*</u>: χ^2^=84.264, p=0.00001, df=14, χ^2^/df=6.02, RMSEA= 0.065, p-close=0.30, SRMR=0.038, CFI=0.94, TLI=0.92) in Table 3. Composite reliability was good, showing estimates of ρ_Anxiety_ = 0.78 and ρ_Depression_ = 0.70.

It is plausible that if one is feeling cheerful, one is improbable to feel anxiety at the same time, and vice versa. Thus, it is theoretically meaningful that feeling cheerful (item 6_D_) and feeling anxiety correlate negatively. Accordingly, in **Model-2** we included a path (cross-loading) from item 6_D_ (“*I feel cheerful”)* to the Anxiety construct, which improved the fit considerably: χ^2^=317.951, p=0.0001, df=75, χ^2^/df=4.24, RMSEA=0.046, p-close=0.262, SRMR=0.042, CFI=0.93, TLI=0.92. However, still the fit was not good.

An extremely high MI between the items 2_D_ (*“I still enjoy the things I used to enjoy”*) and 12_D_ (*“I look forward with enjoyment to things*”) was uncovered. It is rational that still enjoying things and looking forward to things with enjoyment correlate. Therefore, we included a correlated error term between item 2_D_ and 12_D_ which further improved the fit in **Model-3**: χ^2^=296.919, p=0.0001, df=74, χ^2^/df=4.01, RMSEA=0.050, p-close=0.452, SRMR=0.040, CFI=0.94, TLI=0.92. For Model-3, composite reliability (ρ_c_) was 0.78 for the anxiety subscale and 0.65 for depression subscale. Still, 14 normalized residuals were significant, asking for several cross-loadings and correlated errors.

Therefore, we checked if a unidimensional solution termed **Model-4** would fit better, though revealing an exceedingly bad fit: χ^2^=840.194, p=0.0001, df=77, χ^2^/df=10.91, RMSEA=0.091, p-close=0.0001, SRMR=0.070, CFI=0.78, TLI=0.74. However, internal consistency was good ρ_HADS_=0.81. Hence, possibly the misspecification was caused by error covariances. Consequently, we turned to Model-3, again scrutinizing the MI values. The items 8_D_, 12_D_ and 14_D_ displayed MIs>15. These items also displayed low loadings indicating poor reliability as indicators for the depression construct. Possibly, removing some of these items would improve the model fit. Nonetheless, composite reliability was ρ_Depression_=0.65; hence, removing items would cause a week construct with even lower reliability.

All the tested models revealed a chi-square indicating misspecification. However, it is well known that chi-square as a model fit index has limitations. First and foremost, chi-square is sensitive to sample size (48). The present sample is large (N=1190). For that reason, we randomly split the data in two equally sized parts with n=595, representing a sample size more suitable for SEM (structural equation modeling) (47-49). We termed the two parts as Sample1 and Sample2 and tested the original HADS version (Model-1) in both samples. Except the chi-square showing better values, the fit indices demonstrated a similar pattern: **Sample1** χ^2^=225.391, p=0.0001, df=76, χ^2^/df=2.97, RMSEA=0.058, p-close=0.074, SRMR=0.057, CFI=0.90, TLI=0.88, ρ_Anxiety_=0.76 and ρ_Depression_=0.67. **Sample2** χ^2^=257.228, p=0.0001, df=76, χ^2^/df=3.38, RMSEA=0.063, p-close=0.005, SRMR=0.053, CFI=0.91, TLI=0.90, ρ_Anxiety_=0.79 and ρ_Depression_=0.73. Loadings ranged between 0.31 and 0.67 for Sample1 and between 0.29 and 0.77 for Sample2.

**Figure 1.**
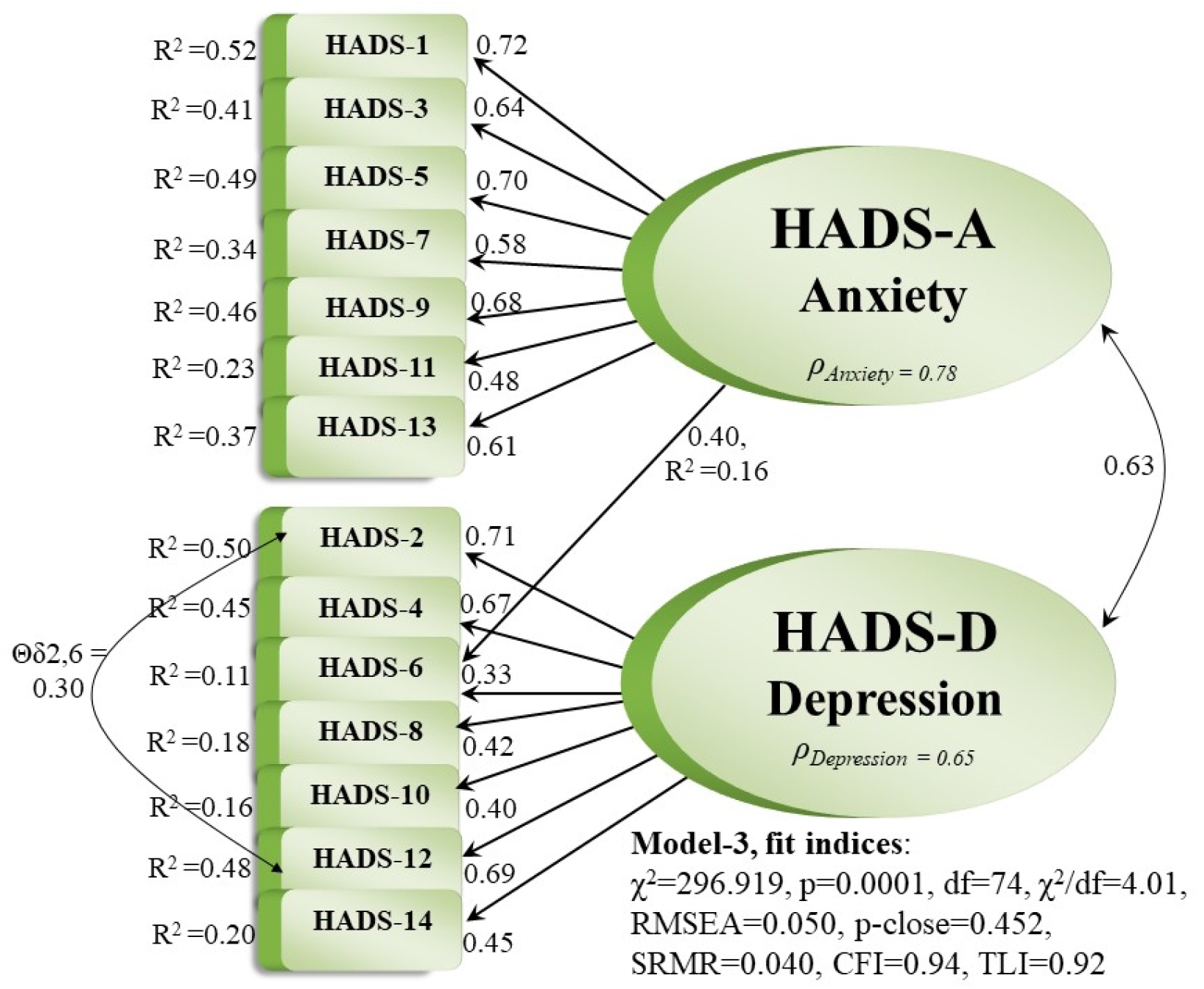
Model 3 – the best fitting measurement model.

Finally, Spearman’s rank correlation was computed to assess the relationship between HADS and overall global QOL (“Thinking about your life at the moment, would you say that you mostly are satisfied with life, or are you mostly dissatisfied?”). There was a negative correlation between high HADS-A score and overall good global QOL, (r=-0.408, p = <0.001) and high HADS-D score and overall good global QOL, (r = -0.450, p = <0.001).

## Discussion

According to the European Commission’s Green paper on mental health (50), depression is one of the most prevalent mental health problems facing European citizens today. The incidence of depression with increasing age is stated (15), simultaneously the number of adults over 80 years is globally expected to dramatically increase in the coming decades (51). Hence, access to a valid and reliable scale assessing anxiety and depression among older community-dwelling adults is highly warranted. Therefore, the aim of the present study was to evaluate the psychometric properties of the HADS among community-dwelling older Norwegians ≥70 years. The present sample included 1190 older adults, with a mean age of 76.5 years. To the authors’ knowledge, no previous studies have examined the psychometric properties of the HADS in a Norwegian population among community-dwelling older adults by means of CFA.

The CFA approach eliminates the need to summate scales because the SEM programs, such as STATA, compute latent construct scores for each respondent. This process allows relationships in the model tested to be automatically corrected for error variance which is a fundamental strength of CFA to construct validation. Thus, the resulting estimates are adjusted for measurement error (47, 49). In this study, the original version of the HADS (Model-1) showed only partly a good fit. Especially, the chi-square demonstrated extremely high values indicating misspecification. However, utilizing the chi-square as a model fit index relates to some limitations. As already stated, chi-square is sensitive to sample size: a misfit may be trivial, still with larger samples the p-value decreases followed by higher estimates (48). This means, that in practice, the chi-square test is “not always the final word in assessing fit” (52). The present sample size is large (N=1190) revealing extraordinarily high estimates for the chi-square. When splitting the file into two parts giving a sample size N=595, the chi-square improved substantially, and the RMSEA was still acceptable. Hence, reflecting on the chi-square statistic in the light of the large sample size, a wide variety of other indices were included to assess model adequacy. The SEM literature states that as a minimum, RMSEA, CFI, and SRMR should be reported in combination with chi-square (46). The use of multiple fit indices provides a more holistic view of goodness of fit, accounting for sample size, model complexity, and other considerations relevant to the study.

Conversely, the RMSEA estimate has demonstrated lower values with large sample sizes (53, 54). For an acceptable fit RMSEA should be ≤ 0.080 (46, 47, 49) or ≤0.10 (42), while estimates ≤ 0.050 suggest a good fit. Looking at Model-1, the RMSEA along with SRMR were acceptable and almost good (0.059, 0.052, respectively), while the CFI and TLI were too low. Concerning CFI and TLI, including a cross-loading item (6_D_) along with a correlated error term between the items 2_D_ and 12_D_ improved these fit indices as well as the total model fit. Consequently, low reliability and content validity seemed to cause the low values for CFI and TLI.

### Dimensionality

Concerning the dimensionality of the HADS, the two-factor-model undoubtedly showed the best fit to the present data; the dimensionality of the HADS questionnaire stood out to be unquestionable. The two factors were properly correlated.

### Reliability

All items were significant. Largely, the items revealed good loadings accompanied by good multiple squared correlations (R^2^) demonstrating good reliability. Nevertheless, particularly three items belonging to the Depression construct (8_D_,10_D_,14_D_) demonstrated low factor loadings and thus poor reliability, explaining very little of the variance in the construct. These three items caused a low reliability coefficient for depression, while anxiety displayed good reliability.

### Construct validity

Construct validity concerns if the set of measured items reflects the theoretical latent construct those items are designed to measure. Hence, it deals with accuracy of measurement involving psychometric evidence of convergent and discriminant validity (55). A measure is said to process convergent validity if independent measures of the same construct converge or are highly correlated (35). Normally, researchers do not have data on two different e.g., depression scales scored by the same sample: this represents a frequent problem connected with convergent validity. However, measures which theoretically are predicted to significantly correlate with depression might be used. The present study included measures of overall global QOL to test for convergent validity, which was supported by a significant correlation in an expected direction.

Discriminant validity entails that a measure must not correlate too highly with other measures from which it is expected to diverge (35). The anxiety and depression factors performed like distinct concepts, supporting the discriminant validity. Simultaneously, the factor correlation between anxiety and depression was highly significant supporting convergent validity (35). The convergent and discriminant validity were further supported by significantly correlations in the predicted direction for anxiety and depression towards QOL.

### ontent validity – a vital aspect of construct validity

Content validity is a central aspect of construct validity. Reliability and content validity represent interrelated measurement properties. In fact, despite a good reliability content validity might be poor. Contrariwise, validity cannot be good if reliability is low (35). The item 8_D_ concerns *“I feel as I’m slowed down”* demonstrated low reliability and thereby poor validity. In the present sample with a mean age of 76.5 years, most individuals have for a long time been outside an active work-life having lots of time to adjust to a slower pace of life. Possibly, ‘feeling slowed down’ does not correspond well to older home-living adults’ daily experiences in relation to depression. This item did not perform to be a valid nor reliable indicator of depression in this population. What is more, about 50% of the participants reported physical or mental long-term illness, injury, or loss of function in daily life. Relevantly, a slower pace of life might seem natural and not necessarily an indicator of depression (2). Likewise, item 10_D_ *“I have lost interest in my appearance”* did not communicate well with these older adults, indicating low reliability and content validity. Losing interest in one’s appearance did not act like a valid indicator of depression in this population. Loose of interest in one’s appearance may be reasoned in the inevitable age-related changes they experience rather than as a depressive symptom. Moreover, item 14_D_ *“I can enjoy a good book or TV program”* also stood out as an unreliable indicator of depression. Plausibly, being old, enjoying a good book or watching TV do not relate with depression. Living in your seventies-eighties-nineties, passive leisure activities are common activities which is useful as restoration time after active leisure activities and related to QOL (56). Reading books might be more demanding due to a declined sight as well as fatigue. Consequently, item 14_D_ did not explain any variance in the depression construct and thus misbehaved as a valid indicator for the depression construct. These findings are consistent with previous studies among nursing home residents without cognitive impairment (28) and in hospitalized older adults (29) where the same three items were troublesome among older adults in Norwegian care facilities. In older ages, for the first time in their life retired adults can slow down. Also, due to a decline in age-related reserve capacity and fear of falling which is the most common fear in older adults (57), many older adults may be forced to a slower pace. Doing passive activities such as watching TV or reading may also be a consequence of having chronic medical condition and multimorbidity which is associated with anxiety and depression (58). Hence, to improve reliability the wording of the items 8_D,_ 10_D_ and 14_D_ should be carefully considered. Furthermore, the former validation study among older adults in nursing homes (28) also involved a cross-loading for item 6_D_ to anxiety, as well as highly significant error variances between items 2_D_ and 12_D_. Surprisingly, home-dwelling older adults (the present study), nursing home residents (two different samples giving an approximate N=500; mean age 84.5 and 86 years) (28) and hospitalized older adults (N=484; mean age 80.7 years) (29) respond similarly findings of the HADS-D items.

Summarized, construct validity and reliability of anxiety were good. Conversely, the depression construct revealed low validity and reliability which are interrelated measurement properties. Exclusively, content validity includes to which extent elements of a measurement scale are appropriate to and characteristic of the specific construct for a certain assessment purpose (35). In this study, content validity concerns if the 14 HADS items and the two-factor dimensionality precisely represent anxiety and depression in this population. Besides, evidence of face validity can be considered as one aspect of content validity (35). High face validity of an instrument increases its use in practical situations via ease of use, proper reading level, clarity, and appropriate response formats. Thus, to improve content validity and thereby also reliability for the depression factor, qualitative studies could be applied to get closer to the actual content of depression, investigating what might be the most essential indicators for depression among community-dwelling older adults. Based in such novel evidence, the three troublesome items could be formulated in a more valid format.

### Strengths and limitations

A notable strength of this research is the empirical examination of the HADS which has not been tested previously in a community-dwelling population in Norway using CFA. Also, the large sample size is a strength, giving the possibility to randomly split the sample into two different samples including 595 community-dwelling older adults each.

Although the older adults were selected randomly in two subsamples, we cannot state that the sample is representative to the community-dwelling older adults in the actual city since 3340 of 5087 declined participation. In addition, those who participated in HUNT4 Trondheim 70+ but excluded from this present study had significantly more often dementia diagnosis, were more often females, had higher age and lower level of education. Hence, we assume that the present sample may be disrupted in that respect not representing all community-dwelling older adults.

## Conclusion

This study showed that the two-factor structure assessing symptoms of anxiety and depression is unquestionable. In conclusion, when we included a cross-loading item (6_D_) along with a correlated error term between item 2_D_ and 12_D_ (Model-3), a good to acceptable measurement reliability was demonstrated, and construct validity was supported.

However, concerning the internal consistency, the original version of HADS (Model-1) revealed a good reliability coefficient for anxiety, but a poor estimate for depression; items 8_D_, 10_D_ and 14_D_ stood out to be unreliable and invalid indicators for depression in this population. Therefore, to be valid indicators of depression among community-dwelling older individuals, these items need to be re-worded informed by qualitative studies exploring symptoms of depression among older adults living at home.

## Data Availability

To protect participants’ privacy, HUNT Research Centre aims to limit storage of data outside HUNT databank and cannot deposit data in open repositories. HUNT databank has precise information on all data exported to different projects and can reproduce these on request. There are no restrictions regarding data export given approval of applications to HUNT Research Centre. For more Information: Åsvold BO, Langhammer A, Rehn TA et al,. Cohort profile update: The HUNT Study, Norway. Int JEpidemiol. Published 17. May 2022.

## Abbreviations

CFA: Confirmative factor analysis
CFI: Comparative Fit Index
CSDS: Clinically significant depressive symptoms
HUNT: The Trøndelag Health Study
MDD: Major depressive disorder
RMSEA: Root Mean Square Error of Approximation
SEM: structural equation modeling
SRMR: Standardized Root Mean Square Residual
MI: Modification indices
TLI: Tucker-Lewis Index

## Acknowledgments

The Trøndelag Health Study (HUNT) is a collaboration between HUNT Research Centre (Faculty of Medicine and Health Sciences, Norwegian University of Science and Technology NTNU), Trøndelag County Council, Central Norway Regional Health Authority, and the Norwegian Institute of Public Health. The authors wish to acknowledge the HUNT Research Centre, Faculty of Medicine and Health Sciences, Norwegian University of Science and Technology and National Advisory Unit for Aging for project management in the HUNT4 Trondheim 70+ survey. Further, we wish to acknowledge Trondheim municipality, students from Norwegian University of Science and Technology for collaborating in the data collection, as well as the older adults who voluntarily participated in the study.

## Authors contribution

Heidi Emly Sivertsen: Data collection - Conceptualization, Methodology, Validation, Formal analysis, Investigation, Resources, Data curation, Writing - original draft, Writing - review & editing. Anne-Sofie Helvik: Conceptualization, Methodology, Validation, Writing - original draft, Writing - review & editing. Linda Gjøra: Data collection - Writing - review & editing. Gørill Haugan: Conceptualization, Methodology, Validation, Formal analysis, Writing - original draft, Writing - review & editing.

## Availability of data and materials

The Trøndelag Health Study (HUNT) has invited persons aged 13 - 100 years to four surveys between 1984 and 2019. Comprehensive data from more than 140,000 persons having participated at least once and biological material from 78,000 persons is collected. The data are stored in HUNT databank and biological material in HUNT biobank. HUNT Research Centre has permission from the Norwegian Data Inspectorate to store and handle these data. The key identification in the data base is the personal identification number given to all Norwegians at birth or immigration, whilst de-identified data are sent to researchers upon approval of a research protocol by the Regional Ethical Committee and HUNT Research Centre. To protect participants’ privacy, HUNT Research Centre aims to limit storage of data outside HUNT databank and cannot deposit data in open repositories. HUNT databank has precise information on all data exported to different projects and can reproduce these on request. There are no restrictions regarding data export given approval of applications to HUNT Research Centre. For more Information: Åsvold BO, Langhammer A, Rehn TA et al,. Cohort profile update: The HUNT Study, Norway. Int JEpidemiol. Published 17. May 2022.

## Declarations

### Ethics approval and consent to participate

All participants received oral and written information and gave informed written consent prior the study. The project was submitted to the Regional Ethics Committee for Medical and Health Research (REC) in Mid-Norway, reference REK 2021/348836, and was approved. This study was conducted in accordance with the ethical principles stated in the Declaration of Helsinki.

